# Measuring plasma volume in women of reproductive age – a comparison study of hydroxyethyl starch to other methods

**DOI:** 10.1101/2025.01.08.25320201

**Authors:** Leigh A. Martin, Kelly Gallagher, Amrita Arcot, Matthew Barberio, Emily R. Smith, Alison D. Gernand

## Abstract

Methods for measuring plasma volume (PV) have rarely been validated or compared, and some are unsafe in certain populations (e.g., pregnancy). We aimed to develop and evaluate a PV method using hydroxyethyl starch (HES) that is safe in women of reproductive age and could be used in pregnancy. A convenience sample of healthy nonpregnant women (n=12) of reproductive age participated in a comparison study using two indicator dilution methods – HES and indocyanine green (ICG), and three estimations - Kaplan, Hurley, and Nadler equations. Baseline blood samples were collected; we injected ICG and HES separately, each followed by post-injection sampling. We compared HES PV to the other methods using Bland-Altman analysis. Participants had mean ± SD age of 25.8±7.5 years and mean BMI of 21.7±1.7 kg/m^2^. Mean PV estimations for HES, ICG, Kaplan, Hurley, and Nadler methods were 2,046±392 mL, 2,765±820 mL, 2,443±464 mL, 2,407±301mL, and 2,373±406 mL, respectively. In each analysis, differences for 11/12 participants were within the Bland-Altman limits of agreement, ±2 SD from each mean difference. In conclusion, PV measured by HES was >300 mL lower than other estimates, but had a narrow distribution within the expected range. Future studies should validate methods for PV measurement across populations, including pregnancy.

**Highlights:**

- What is the central question of this study? How does plasma volume (PV) estimated using hydroxyethyl starch (HES) compare to estimates using other methods in healthy women of reproductive age?
- What is the main finding and its importance? In this comparison study of PV methods (HES, indocyanine green, and estimation equations), PV measured by HES was >300 mL lower than other estimates, but had a narrow distribution that was within the expected range. Future work is still needed to validate the HES method across populations.

## Introduction

In a healthy individual of normal physiologic status, plasma accounts for approximately 60% of whole blood, with the remaining 40% consisting of red blood cells and other blood components (1). Previous studies have reported that nonpregnant females have mean PV values of approximately 2,500 mL to 2,600 mL (2,3). Plasma is important for coagulation, immune defense, maintenance of osmotic pressure, thermoregulation, and transport of nutrients (4). Plasma volume (PV), as a component of whole blood, can change because of both normal and abnormal conditions, resulting in hypovolemia (low blood volume) or hypervolemia (high blood volume). For example, intense exercise has been found to increase PV due to the temporarily increased plasma albumin concentration (5). Dehydration has been shown to decrease PV after periods of heat exposure (6). In pregnancy and other physiologic states, plasma increases proportionally more than red blood cells, resulting in hemodilution. Most micronutrient biomarkers are measured in plasma (or serum), but rarely is PV measured to assess the potential relationship.

There are two general categories for the methods of PV estimation: direct laboratory-based dilution methods and indirect estimation equations (7,8). All laboratory-based methods, including radiolabeled molecules (iodinated human serum albumin) and dye dilution methods (Evans’ blue and indocyanine green (ICG)), follow an indicator dilution technique. Carbon monoxide rebreathing and hydroxyethyl starch (HES) are other non-dye, laboratory-based dilution methods that have been used in past studies (9,10). The laboratory-based methods have been shown to be more accurate compared to estimation equations, but they can include exposure to radioactive materials, be potentially invasive, or require expensive and technologically advanced laboratory methods or equipment. Due to these disadvantages, they also may not be acceptable or safe to use in vulnerable populations, like pregnant individuals (7,8).

There are several equations that are used to estimate blood volume, red cell volume, and PV. These equations use health data that is often readily available, such as age, height, and weight; some equations utilize body surface area (BSA), which is calculated using a validated equation and an individual’s weight and height (11). These equations are easy to use and non-invasive, but they may not be as accurate as laboratory-based methods that more directly measure PV (7,8,12).

There have been many calls for additional research in PV, with emphasis to develop methods for special populations (including pregnant individuals) and to estimate PV in various settings and/or disease states (8,13). This study aimed to develop and evaluate a method to measure PV using HES in healthy women of reproductive age. Within this aim, we compared PV measured by HES to PV measured by ICG and PV estimated by equations. We hypothesized that the HES method would produce similar results to the other tested and validated methods, each within a 10% difference.

## Methods

We conducted a pilot comparison study to develop and evaluate a method to measure PV using HES in healthy women of reproductive age. This study was part of the protocol development stage of a single-blinded, stratified, multiple ascending dose trial for vitamin B12 supplementation during pregnancy in Bagamoyo, Tanzania (https://clinicaltrials.gov/study/NCT05426395) in which PV was measured using HES.

### Sample Size and Eligibility Criteria

We aimed to recruit a minimum of 10 participants. Eligible individuals were non-pregnant females aged 18 to 44 years in general good health (did not have a known, ongoing health condition or medical issue that required regular monitoring by a doctor or regular visits to the hospital) that had a BMI of 18.5 to 24.9 kg/m^2^, were non-smokers, and had at least 12 months since their last pregnancy if applicable. Individuals were ineligible if they had any of the following: 1) known allergy to corn, HES, shellfish, or iodine; 2) low or high blood pressure on the day of measurements (SBP <90 or ≥130 mmHg and/or DBP <60 or ≥80 mmHg); 3) self-reported low or high blood pressure (SBP <90 or ≥130 mmHg and/or DBP <60 or ≥80 mmHg) (14); 4) current hypertension or previous hypertensive disorder in pregnancy (such as gestational hypertension); 5) taking regular medication prescribed by a doctor for a health condition, other than hormonal contraception; 6) polycystic ovarian syndrome, or renal, liver, autoimmune, or bleeding disorders; 7) congestive heart failure; 8) pregnant or currently trying to conceive; 9) currently breastfeeding; and 10) self-reported history of difficult blood draws or IVs. The study protocol was approved by the Institutional Review Board of the Office for Research Protections at The Pennsylvania State University (STUDY00016189) and participants signed informed consent at their study visit.

### Study Procedures

In this cross-sectional comparison study, a convenience sample of eligible participants were recruited between September 2021 and July 2022 via paper fliers and various electronic methods (StudyFinder, ResearchMatch, FIRSt Families) in State College, Pennsylvania, and the surrounding areas within Centre County, Pennsylvania. Interested individuals completed a phone pre-screening; if deemed provisionally eligible based on the pre-screening, individuals were scheduled to visit the Penn State Clinical Research Center (CRC) for enrollment into the study and to complete an in-person screening.

Prior to the study visit, participants were instructed to fast (no food or beverages except water) for 12 hours and to refrain from alcohol consumption for 48 hours prior to the visit. We recommended that participants drank approximately 8 cups of water per day, starting 48 hours prior to the visit, to ensure adequate hydration. They were also instructed to void the first morning urine and we recommended that they consume an additional 1-2 cups of water on the morning of their visit. At the end of the in-person screening, we asked participants if they adhered to the fasting requirements; if they reported food or alcohol consumption within the last 12 or 48 hours, respectively, we ended their visit and rescheduled for the next available day. We did not ask participants about their adherence to our recommended water consumption.

At the start of the visit, participants provided voluntary informed consent and were enrolled into the study; enrollment at this point was needed to complete the in-person eligibility screening and the remainder of the visit. They then provided a urine sample, which was used to conduct a pregnancy test (QuickVue, Quidel Corporation, San Diego, CA, USA) as part of the in-person screening. Participants were weighed to the nearest 0.1 kg using a digital scale, and their height was measured to the nearest 0.1 cm using a portable stadiometer; body mass index (BMI: kg/m^2^) was calculated with these measurements. Both weight and height were measured once. Blood pressure and pulse were each measured twice following the American Heart Association protocol, starting after 5 minutes of undisturbed rest using an electronic blood pressure monitor (OMRON HEM-712C Blood Pressure Measuring Machine). Mean systolic and diastolic blood pressure were used for eligibility (14). After the in-person screening, ineligible participants did not continue with the remaining study visit activities; they were compensated $50 cash.

If eligible based on the in-person screening, trained study staff administered a questionnaire to collect demographic, health history, and pregnancy history data. Body fat percentage was then measured using a segmental bioelectric impedance analyzer (Tanita BC534 Glass InnerScan Body Composition Monitor, Tanita Corporation of America Inc., Arlington Heights, IL). Hydration status was assessed using the urine sample and a urine specific gravity pen refractometer (Atago 3741 PEN-Urine S.G. Digital Handheld Pen-Style Refractometer, Atago USA, Inc.); hydration status was categorized as hydrated (1.000-1.019), moderately dehydrated (1.020-1.027), or severely dehydrated (1.028-1.035) (15).

Participants were then asked to lay on their left side and rest undisturbed in a clinic room for 15 minutes in preparation for establishment of the intravenous (IV) catheter in the left arm. A heating pad was placed on the left arm during the rest period. We selected this position to align with standard practices for blood sample collection during pregnancy, as we developed the HES method to be used in pregnancy. A trained nurse inserted the IV; collected baseline blood samples into vacutainers (6 mL trace element-free EDTA-free vacutainer for serum, 6 mL trace element-free K2EDTA vacutainer for plasma, and 2 mL K2EDTA vacutainer for whole blood; BD Vacutainer, BD, New Jersey, USA); injected the ICG and HES solutions; and collected post-injection blood samples into vacutainers (ICG and HES procedures are detailed below). We injected the ICG solution first followed by the HES solution because HES is used clinically as a volume expander, and we did not want the HES volume to influence the PV estimated by ICG. After the IV was removed, the participants were observed for approximately 15 minutes (to ensure no allergic reaction and overall feeling well), during which we measured blood pressure and pulse twice and provided $50 cash as a stipend for participation.

### Indocyanine Green (ICG)

After the establishment of the IV catheter and collection of baseline blood samples, the syringe used for the ICG solution injection was weighed using a high-precision scale before the solution was prepared, after the syringe was filled, and then again after the solution was injected; these weights were later used to calculate how much ICG was injected. The solution (IC-Green, AKORN Inc, Lake Forest, IL, USA) was prepared by the nurse manager as 0.25 mg/kg body weight, which was measured at the start of the visit as part of the in-person screening. The nurse then injected the solution as a bolus dose over 5 seconds. Once the injection was complete, the nurse collected 3 mL K2EDTA vacutainers (BD Vacutainer, BD, New Jersey, USA) for plasma, starting at 2 minutes and occurring every 45 seconds, up to 5 minutes (a total of 5 blood draws at exactly (min:sec) 2:00, 2:45, 3:30, 4:15, and 5:00) (16). Plasma samples were centrifuged at 3,200 r/min for 15 minutes at room temperature, and we aliquoted the plasma into cryovials for PV measurement.

Plasma volume was measured using the indicator-dilution principle; baseline plasma (called the “blank” plasma sample) and the 5 post-injection samples were used to estimate PV. The blank plasma sample, prepared standard solutions, and post-injection samples were each loaded into a 96-well plate in triplicate and read at a wavelength of 805 nm on an Epoch plate reader (BioTek Instruments, Inc.) powered by Gen5 Software. We then constructed a standard curve of absorbance against the standard concentrations; this was used to estimate the ICG concentrations of the 5 post-injection plasma samples. We back-extrapolated the ICG concentration to the time of injection (t = 0 seconds) because of its rapid hepatic clearance and calculated PV for each participant as:

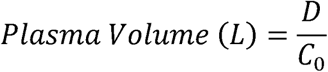

where D was the dose of ICG administered in mg and C_0_ was the plasma concentration of ICG in mg/L at t = 0 seconds. Further details on the PV determination have previously been published (16).

### Hydroxyethyl Starch (HES)

We developed a method to measure PV using HES by adapting previously published methodology (10,12). Immediately following the post-ICG injection sample collection, the trained nurse injected a 120-170 mL bolus of HES (6% hetastarch in 0.9% sodium chloride injection, average molecular weight: 600,000; Hespan, B. Braun Medical Inc.) through the IV catheter over 4 minutes. Ten participants received 120 mL, and 2 participants received 170 mL of HES; we tested different volumes of HES as part of our method development. The timer was reset and the nurse collected 3 mL post-HES injection blood samples into K2EDTA vacutainers (BD Vacutainer, BD, New Jersey, USA). We collected samples at 5, 10, and 15 minutes after the injection as part of our method development, to confirm the timing of collection in the previous study (10). The samples were kept at room temperature and centrifuged in the same way as the ICG samples, and plasma was aliquoted into cryovials for PV determination. Due to glycolysis that could continue to occur in whole blood, it is ideal to process samples within 30 minutes of collection; however, we did not see the timing mentioned in other protocols and did not take this timing issue into consideration (we now recommend doing so). We estimated that plasma samples were processed within 60 minutes of collection.

Post-HES injection plasma samples were transported to the lab in a cooler with an icepack, and we immediately assayed them before being frozen for storage. The whole blood samples were assayed for hematocrit, using a HemoPoint H2 Analyzer (EKF Diagnostics, USA). If a participant was anemic based on hemoglobin assay (HemoCue Hb801, HemoCue AB, Sweden), we measured hematocrit using a microhematocrit centrifuge (ZipCombo Centrifuge, LW Scientific, USA) because the analyzer would not measure anemic values – this was done by drawing whole blood into a capillary tube, centrifuging, then measuring the length of the column of the packed red cells with a hematocrit reader/ruler. Hematocrit values were used in the PV equation.

For the plasma processing, we first transferred 0.6 mL of each plasma sample to a cryovial, then added 0.15 mL of 12M hydrochloric acid to each sample and vortexed thoroughly. The cryovials were placed in a boiling water bath for 7 minutes and then in a room-temperature water bath for 2 minutes. The acid and water bath were done to hydrolyze HES in the samples. The samples were then vortexed, and 0.65 mL of tris buffer was added to each, followed by additional vortex mixing and benchtop incubation for 6 minutes, to neutralize and stop the hydrolysis reaction. After incubation, the samples were vortexed again and the pH was measured using pH strips; the target pH was 7.0 +/- 0.5, and additional tris buffer was added to adjust the pH if needed. The samples were then centrifuged at 3600 rpm for 16 minutes, and the supernatant was obtained and assayed for glucose using a handheld glucometer (HemoCue Glucose 201, HemoCue AB, Sweden).

We calculated blood volume at baseline and 5-, 10-, and 15-minutes post HES injection according to the equation below.

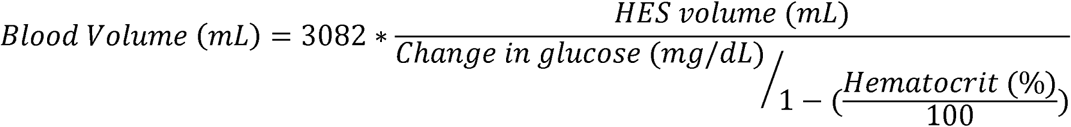

Hematocrit (%) was determined from the baseline whole blood sample as described above, and change in glucose (mg/dL) is equal to the post-HES injection glucose reading minus the baseline glucose reading (10,12). The constant factor, 3082, was determined by Tschaikowsky *et al.* using a sample of 50 healthy volunteers, and we used this factor in our equation given our similarly healthy population (12). We then converted blood volume to PV using the following equation:

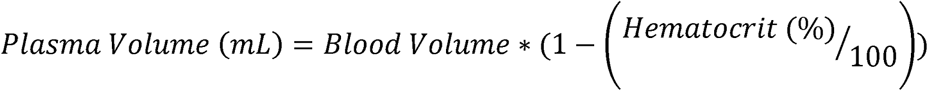

The original study conducted by Tschaikowsky *et al.* used a solution of 10% HES (12), but we purchased a solution of 6% HES (10% not available in the United States). Due to the concentration difference, we converted the 100 mL volume of HES injected in the Tschaikowsky study to the weight in grams of HES based on the original product. We then converted the grams back to the equivalent volume of a 10% HES solution (102 mL converted to 120 mL; 72 mL converted to 170 mL) and used that value in the equation. We tested 120 mL and 170 mL as part of our method development because the lower volume was easier to inject (in two 60 cc syringes) compared to 170 mL, but 170 mL was closer to the original HES validation study.

We calculated PV estimates with the three collection timepoints (5, 10, and 15 minutes) as part of our method development. In general, the estimate at 5 minutes post-injection was higher than the other timepoints, and the estimate at 10 minutes post-injection produced results within normal expected ranges. The estimate at 15 minutes post-injection was overall similar to the 10-minute timepoint. For comparison of PV measurements between methods, we used 10 minutes post-HES injection as this time was used by previous studies (10,12).

### Estimation Equations

There are over a dozen published equations to estimate PV or blood volume (7). We selected equations that included at least two variables collected as part of our study, which narrowed the options since some equations use only one datapoint (e.g., weight). As well, we chose equations that were easy to implement clinically, using data that would be simple to collect or already in an individual’s health record. We did not consider the accuracy or validation for each chosen equation, as these data were not always easily accessible in publications.

### Kaplan Equation

Kaplan’s equation for was used to estimate PV. The equation is (17):

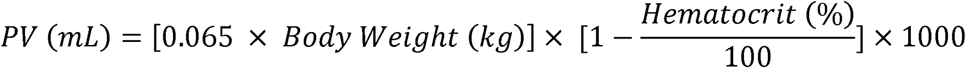

The participant’s body weight was measured at the start of the visit, and the hematocrit was measured from the baseline whole blood sample.

### Hurley Equation

We used Hurley’s equation to estimate PV. The equation is (18):

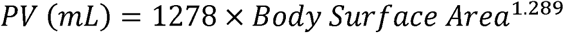

Body surface area (BSA) was calculated for each participant using the following equation (11):

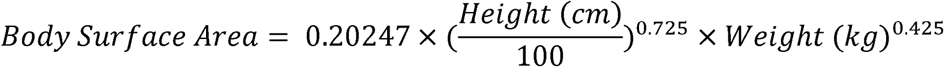

### Nadler Equation

Nadler’s equation was used to estimate total blood volume which we then converted to PV. The equation is (19):

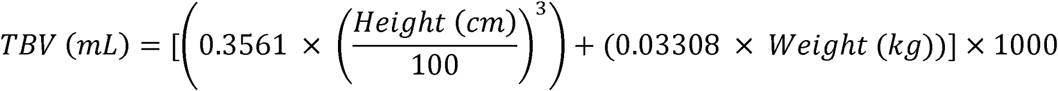

Plasma volume was estimated by converting TBV to PV using the conversion equation above.

### Statistical Analysis

Categorial variables were presented as frequencies (%). Normality of continuous variables was assessed by testing the distribution of each variable against a normal distribution using the Shapiro-Wilk test and inspection of kernel density plots. Continuous variables were all normally distributed and were therefore summarized with mean ± SD.

Mean PV values and differences were compared using Bland-Altman analyses for HES at 10 minutes, ICG at t = 0 seconds, the Kaplan PV equation, the Hurley PV equation, and the converted Nadler PV equation. The Bland-Altman analysis is used to assess agreement between a validated method and a novel method. The analysis includes a plot, which shows the mean difference between the two compared methods and a lower and upper limit of agreement (LOA). The lower LOA is placed -2 SD away from the mean difference, and the upper LOA is placed +2 SD away. If the methods are in perfect agreement, the mean difference line would be at zero (20). Statistical analyses were conducted in R (version 4.1.1).

## Results

Seventeen participants were enrolled in the study, and a total of 12 participants had complete visit data and were included in the final analysis. Mean ± SD age of participants was 25.8 ± 7.5 years with BMI of 21.7 ± 1.7 kg/m^2^ (**Table 1**). They were predominantly white, non-Hispanic, college-educated, and never married. Over 80% of participants reported middle/average income; no participants reported that their income was low. All but one of the participants were nulliparous, and 58% reported using contraceptives.

**Table 1.** Characteristics of study participants (healthy women of reproductive age) (n=12).

| Participant Characteristics | Mean (SD) or n (%) |
| --- | --- |
| Age, years | 25.8 (7.5) |
| BMI, kg/m<sup>2</sup> | 21.7 (1.7) |
| Body surface area, m<sup>2</sup> <sup>a</sup> | 1.64 (0.16) |
| Systolic blood pressure, mm Hg | 110.0 (10.3) |
| Diastolic blood pressure, mm Hg | 72.1 (5.7) |
| Body fat percentage, % | 25.8 (5.6) |
| Urine specific gravity | 1.0161 (0.0075) |
| Urine specific gravity, n (%) <sup>b</sup> |  |
| Hydrated | 8 (67) |
| Moderately dehydrated | 3 (25) |
| Severely dehydrated | 1 (8) |
| Race/Ethnicity, n (%) |  |
| White, Non-Hispanic | 7 (58) |
| White, Hispanic | 1 (8) |
| Black, Non-Hispanic | 1 (8) |
| Asian, Non-Hispanic | 3 (25) |
| Education, n (%) |  |
| High school | 4 (33) |
| Associate/Bachelor's degree | 4 (33) |
| Master's/Doctorate degree | 4 (33) |
| Marital status, n (%) |  |
| Never married | 9 (75) |
| Married | 3 (25) |
| Income level, n (%) <sup>c</sup> |  |
| Middle/Average | 10 (83) |
| High | 2 (17) |
| Contraceptive use, n (%) | 7 (58) |
| Nulliparous, n (%) | 11 (92) |

<sup>a</sup> BSA=0.20247\*(height (cm)/100)<sup>0.725</sup> \*weight (kg)<sup>0.425</sup> (11).
<sup>b</sup> Hydration cutoff values: 1.000-1.019 (Hydrated), 1.020-1.027 (Moderately dehydrated), 1.028-1.035 (Severely dehydrated) (15).
<sup>c</sup> No participants reported that their average income was low.

The mean ± SD PV estimations were 2,046 ± 392 mL (HES), 2,765 ± 820 mL (ICG), 2,443 ± 464 mL (Kaplan), 2,407 ± 301 mL (Hurley), and 2,373 ± 406 mL (Nadler) (**Table 2**). Individual participant estimations for each method are also included in Table 2. **Figure 1** shows the comparison of PV estimations across all methods used in this study. HES had the lowest mean and ICG had the highest mean; HES, Hurley, and Nadler had the lowest SDs and ICG had the highest SD.

**Figure 1.**
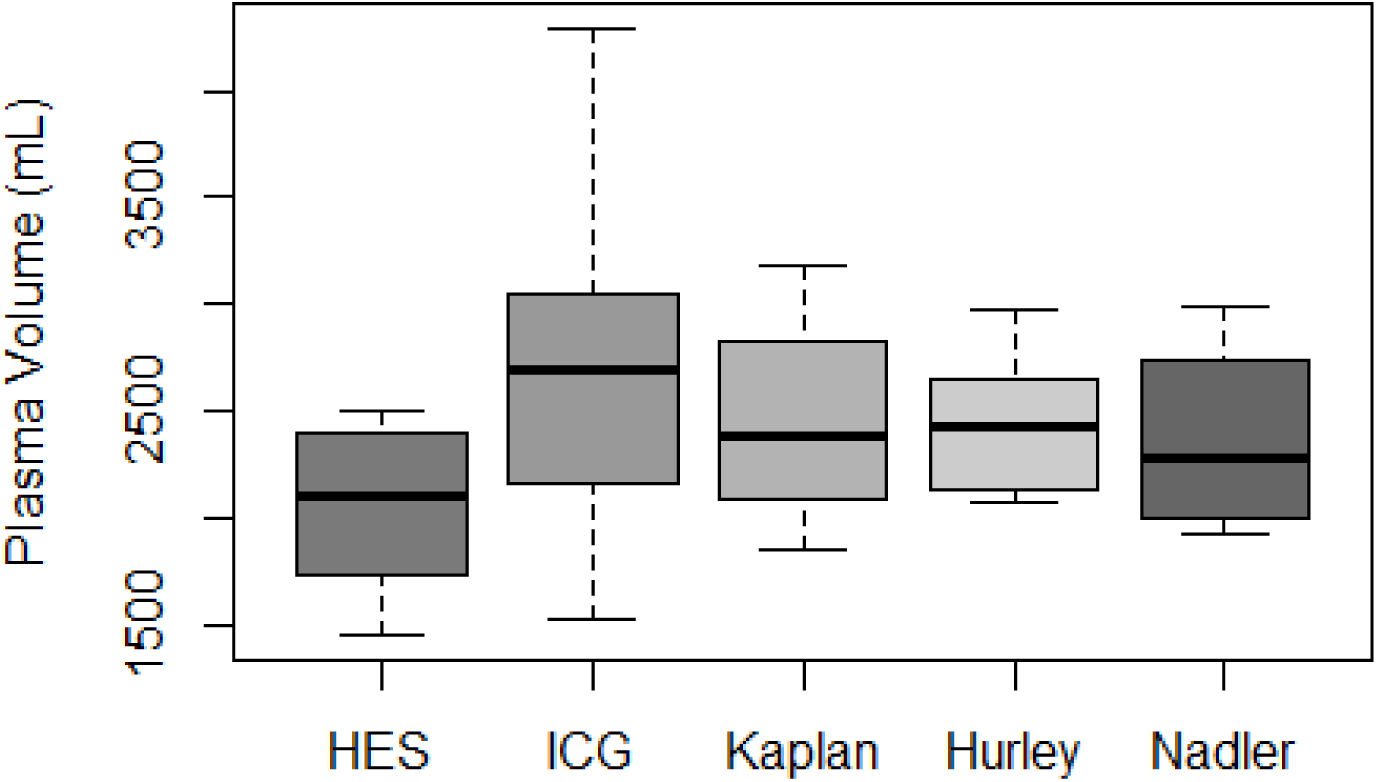
Comparison of PV between laboratory-based methods (HES and ICG) and estimation equations (Kaplan, Hurley, and Nadler) (n=12). Abbreviations: PV, plasma volume; HES, hydroxyethyl starch; ICG, indocyanine green.

**Table 2:** PV across all methods for each participant (n=12).

| Participant number<sup>a</sup> | HES (mL) | ICG (mL) | Kaplan (mL) | Hurley (mL) | Nadler (mL) |
| --- | --- | --- | --- | --- | --- |
| 1 | 1737 | 1991 | 2092 | 2110 | 2062 |
| 2 | 2344 | 3116 | 3127 | 2640 | 2978 |
| 3 | 2495 | 2661 | 3184 | 2748 | 2929 |
| 4 | 2389 | 2723 | 1949 | 2148 | 1945 |
| 5 | 1741 | 2965 | 2323 | 2379 | 2238 |
| 6 | 1503 | 1940 | 1848 | 2078 | 1924 |
| 7 | 2393 | 2864 | 2448 | 2468 | 2419 |
| 8 | 2346 | 4290 | 3062 | 2967 | 2991 |
| 9 | 1454 | 1527 | 2079 | 2155 | 1947 |
| 10 | 1809 | 4133 | 2598 | 2656 | 2538 |
| 11<sup>b</sup> | 2431 | 2328 | 2177 | 2066 | 2170 |
| 12<sup>b</sup> | 1833 | 2637 | 2434 | 2474 | 2335 |
| Mean (SD) | 2046 (392) | 2765 (820) | 2443 (464) | 2407 (301) | 2373 (406) |

Abbreviations: HES, hydroxyethyl starch; ICG, indocyanine green.
<sup>a</sup> The participant number in this column is not the participant's study identifier code.
<sup>b</sup> These participants received 170 mL of HES; all other participants received 120 mL of HES.

**Figure 2** shows the Bland-Altman comparison of the HES PV estimation to the ICG PV estimation. The mean ± SD difference in mL (%), upper, and lower LOA for the comparison was -718 ± 748 mL (28.0%), 748 mL, and -2,185 mL, respectively. **Figure 3** shows the Bland-Altman comparison of the HES PV estimation to the results produced from each estimation equation. The mean ± SD difference in mL (%), upper, and lower LOA for each comparison are as follows, respectively: HES versus Kaplan (-397 ± 410 mL (23.1%), 407 mL, -1,201 mL); HES versus Hurley (-361 ± 380 mL (21.8%), 383 mL, -1,106 mL); HES versus Nadler (-327 ± 367 mL (20.6%), 392 mL, -1,046 mL). The mean difference for each HES comparison with the estimation equations was smaller than the HES versus ICG comparison.

**Figure 2.**
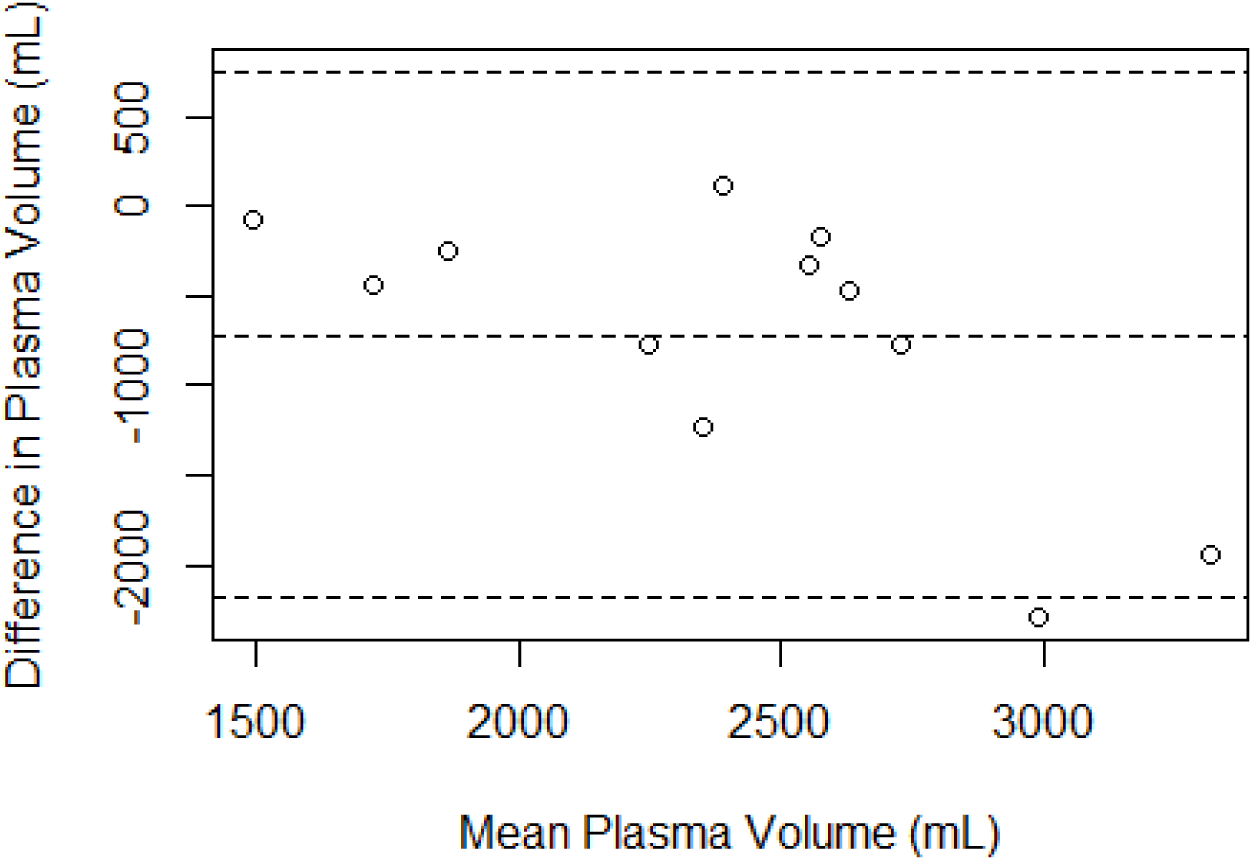
Bland Altman analysis plot for PV comparison between HES and ICG (n=12). The open circles represent each participant’s difference in PV (mL) between the compared methods (HES minus ICG). The middle dashed line represents the mean difference in PV (mL) between the compared methods. The top dashed line represents the upper limit of agreement (LOA), placed +2 SD from the mean difference. The bottom dashed line represents the LOA, placed -2 SD from the mean difference. Abbreviations: PV, plasma volume; HES, hydroxyethyl starch; ICG, indocyanine green.

**Figure 3.**
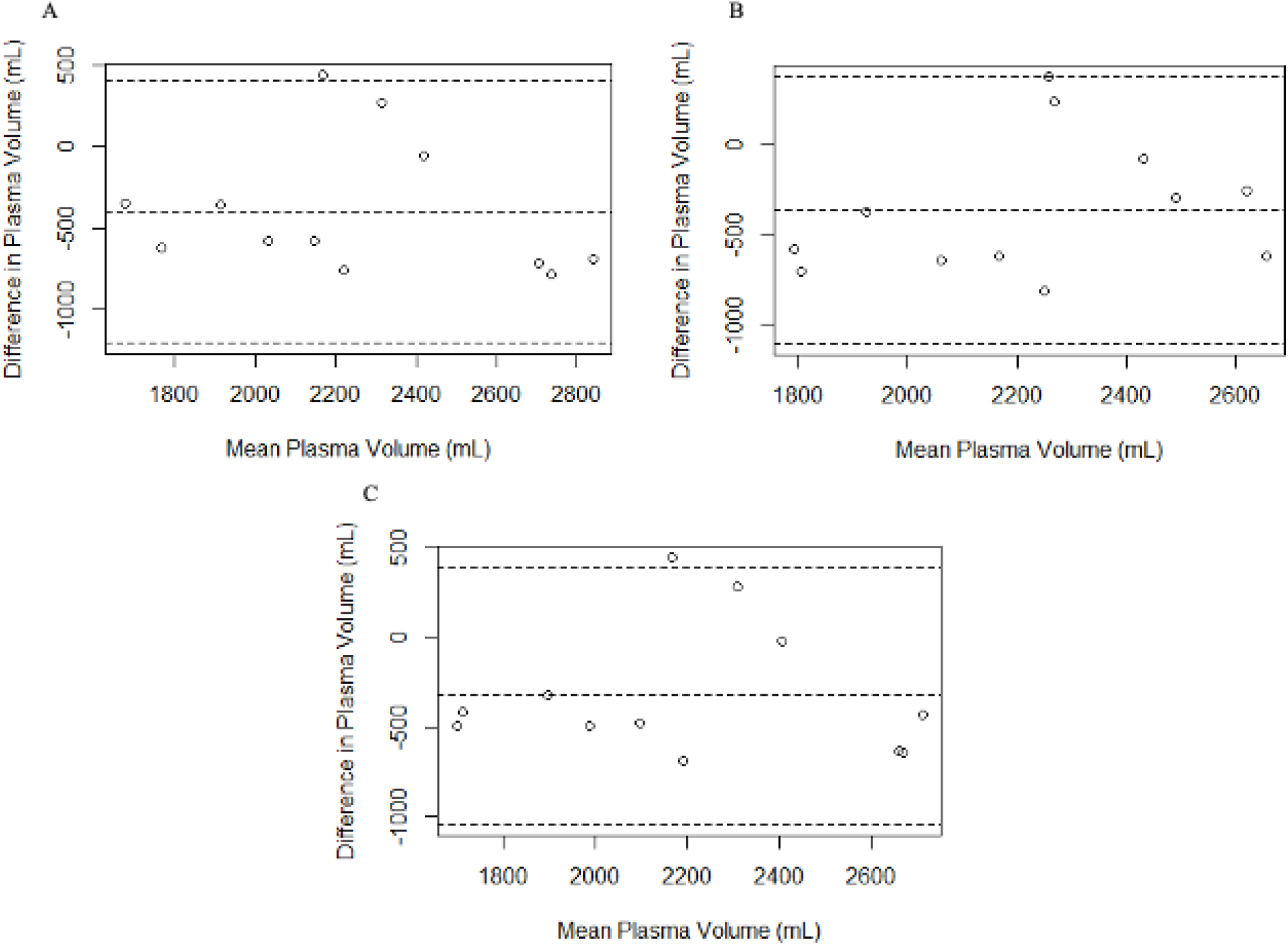
Bland Altman analysis plots for PV comparison between HES and A) Kaplan; B) Hurley; and C) Nadler estimation equations (n=12). The open circles represent each participant’s difference in PV (mL) between the compared methods (A: HES minus Kaplan; B: HES minus Hurley; C: HES minus Nadler). The middle dashed line represents the mean difference in PV (mL) between the compared methods. The top dashed line represents the upper limit of agreement (LOA), placed +2 SD from the mean difference. The bottom dashed line represents the LOA, placed -2 SD from the mean difference. Abbreviations: PV, plasma volume; HES, hydroxyethyl starch.

### Sensitivity Analysis

Ten participants received 120 mL of HES, and 2 participants received 170 mL of HES for the injection. We conducted a sensitivity analysis by excluding the 2 participants who received 170 mL and results were similar.

## Discussion and Conclusions

In this comparison study in healthy women of reproductive age, we sought to develop a method to measure PV that was comparable to another validated method. Our HES method measured PV in a range expected for women of reproductive age. We found that, contrary to our hypothesis (<10% difference in methods), mean HES estimated volume was >20% different from each other method’s mean. The mean HES estimates were the lowest of all methods and were closer to the equation estimates (∼300-400 mL difference) than those of ICG (∼700 mL difference).

To our knowledge, only 2 studies have been conducted to compare HES to other methods for measuring blood volume. Tschaikowsky *et al*. compared the HES method to carbon monoxide rebreathing and an estimation equation in male and female hospital patients undergoing neurosurgery (n = 12) (12). In their study, there was a high level of agreement between the laboratory methods, with a mean difference of 1%, indicating that the HES method could be used in place of the carbon monoxide method to estimate blood volume. The mean difference in blood volume between the laboratory methods was 52.3 ± 183 mL and the limits of agreement (mean difference +/- 2 SD) were -313.7 and +418.3 mL (compared to our -2,185 mL and +748 mL limits of agreement between HES and ICG). However, there was poor agreement between their HES method and the predicted blood volume by the estimation equation, similar to our results.

Vricella *et al*. compared HES to a weight-based estimation equation developed by Feldschuh and Enson to measure blood volume in pregnant women with normal weight (n = 29); the study also included pregnant women with obesity (10,21). Their study found that the HES method produced a consistently higher mean blood volume compared to the estimation equation for participants with normal weight (6,944 ± 2,830 mL versus 4,417 ± 436 mL, respectively) (10). Our study found the opposite, where the HES method produced lower mean PV values compared to our selected estimation equations. This may be partially explained by using a pregnant vs. non-pregnant population, but also the differences in parameters for each equation (10). We did not collect information on deviation from desired weight and were unable to use the same equation as Vricella *et al*.

There has been little other research conducted to examine PV in healthy women of reproductive age. Aguree and Gernand conducted a pilot study to examine the association between PV measured by ICG and micronutrient biomarkers in healthy women of reproductive age (n = 9) (22). The mean PV was 2,067 ± 470 mL in this study, which is close to our mean PV estimation using HES (2,046 ± 392 mL). In another study conducted by Aguree *et al.*, ICG was used to examine PV variation across the menstrual cycle in healthy women of reproductive age (n = 45) (2). The mean PV estimations were 2,276 ± 478 mL (early follicular phase), 2,232 ± 509 mL (late follicular phase), and 2,228 ± 502 mL (midluteal phase); these mean values were ∼10% higher than our HES estimations but ∼20% lower than our ICG estimations. Notably, the standard deviation for our ICG values was much higher than the Aguree *et al.* study (820 mL versus ∼500 mL) (2). We are uncertain why the results of our ICG estimates were so different from our previous ICG work – we followed the same procedures in the same clinic and laboratory. If ICG estimates in the current pilot study were consistent with our prior ICG studies, HES and ICG would have closely aligned, and our hypothesis would have been supported.

In exploring other potential reasons for differences between HES and ICG, we considered the timing of the injections. We chose to inject the ICG solution first (before HES), so that the HES would not be able to influence PV estimated by ICG, however this put our pre-HES blood draw about 10 minutes earlier than it would have been if taken directly before HES injection. We tested glucose values across this interval (n=1) and they were on average within 2 mg/dL of each other. Also, we do not expect that the small amount of HES injected (120-170 mL compared to 1000-1,500 mL used clinically to expand PV) was likely to change plasma volume during the measurements. Finally, we considered the potential effect of glycolysis that may have occurred in the blood tubes before the plasma was separated. We typically centrifuged the samples within 30-60 minutes of blood collection, but we did not have a specific time limit in our protocol. We also used EDTA tubes rather than tubes containing sodium fluoride. Other HES studies also used EDTA tubes and our own testing by hour across four hours did not find consistent differences between glucose values from EDTA vs. sodium fluoride tubes. We plan to conduct additional testing related to glycolysis and effective glycolytic inhibitors in vacutainers.

Other studies have been conducted to compare laboratory methods for measuring PV. Poulsen *et al*. compared PV measured by carbon monoxide rebreathing and Evans’ blue dye in healthy male participants (n = 10) exposed to sea level and high altitude (23). They found that PV measured by carbon monoxide rebreathing decreased by almost 10% at high altitude compared to sea level, while PV measured by Evans’ blue dye did not change. The mean difference between the methods was significantly different from zero in those who experienced hypoxia due to the elevation change; the mean difference at sea level was 0.11 L, and the mean difference at high altitude was 0.43 L. The mean PV estimates using Evans’ blue and carbon monoxide were 3.49 L and 3.39 L, respectively (23). In another study measuring PV in healthy male volunteers, Sawka *et al.* found a mean PV of 3.25 ± 0.41 L, with a range of 2.70 to 4.28 L (24). Menth-Meier *et al*. compared PV measured by ICG and radiolabeled iodinated human serum albumin in both healthy (n = 10) and post-operative (n = 21) adult female and male participants (25). The mean PV measured by ICG produced results closely aligned with PV measured by radiolabeled iodinated human serum albumin; the maximum lack of agreement was -9.1 mL/kg body weight, with all datapoints contained within the lower and upper limits of agreement and the mean difference between methods being close to zero (25).

A strength of our study is the comparison nature; there are few studies that have compared PV methods to examine their agreement and consistency, and there is even less work comparing the HES method to other methods of PV measurement. Unique to past studies, we examined both laboratory-based (HES and ICG) and estimation equation (Kaplan, Hurley, Nadler) methods to measure PV. Additionally, we measured PV in healthy volunteers, while past work has largely focused on critically ill or hospitalized participants, allowing us to provide valuable information on PV in healthy individuals.

However, we did not include the gold-standard radiolabeled iodinated human serum albumin method for measuring PV. Instead, we compared the HES method to a validated method that we have used in previous research (2, 14, 21). We could not find any specific reasons for the differences between our PV estimations and others; we did troubleshooting with the laboratory work and closely reviewed parameters that may have had an influence in the PV estimations, such as height and body fat percentage. In our study, participants were instructed to rest lying down for 15 minutes before the IV was established. It is possible that the ICG results would have been more consistent and stable if participants had been lying down for a longer period of time before the IV was established, such as 30 minutes.

In a small sample of healthy women of reproductive age, our results for the HES method aligned with expected PV estimates in healthy women of reproductive age, but they were lower than the PV estimates of the other methods chosen for this study. It is unclear if results would be consistent across different populations. Overall, the HES method is easy to implement and use, and it is safe for use in a variety of populations, including pregnancy. However, the HES method needs further testing and validation. Additionally, more PV research is needed overall, to understand normal volumes in healthy individuals and disease states, especially in the context of pregnancy.

## Data Availability

All data produced in the present study are available upon reasonable request to the authors.

## Acknowledgments

We would first like to thank the volunteers for their participation in our study. We also thank the medical staff and physicians, especially Cynthia Flanagan and Christa Oelhaf, at the Clinical Research Center of The Pennsylvania State University’s Clinical and Translational Science Institute. As well, we thank Dr. Lacy Alexander in the Department of Kinesiology at The Pennsylvania State University for providing the high-precision scale for the ICG method. Special thanks go to Drs. Sixtus Aguree, Laura Vricella, and Rachel Walker for their guidance and assistance during method development and data troubleshooting. Finally, we sincerely thank Casey Ostrowski for their assistance with participant recruitment.

## Statement of authors’ contributions to manuscript

All authors contributed to critical reading and revision of the manuscript. LAM: study conception, execution, laboratory analysis, data analysis, interpretation, and manuscript writing. KG: study conception, execution, and interpretation. AA: study execution and interpretation. MB: study conception and interpretation. ERS: study conception and interpretation. ADG: study conception, execution, and interpretation.

